# Identification and Multivariate Analysis of 16 Compositional Elements of Relaxation—A Web-based Music Listening Study

**DOI:** 10.1101/2023.08.09.23293385

**Authors:** Claudius Conrad, Eric Nathan, Christoph Wagner, Susan Xu, Todd Frazier, Mei Rui

## Abstract

**Background:** Music is an effective non-pharmacologic, non-invasive, safe, and low-cost intervention to enhance psychophysiological wellness and promote relaxation. This study addresses major knowledge gaps in establishing and validating a scientifically reproducible and rigorous methodology for music repertoire selection to enhance perceived relaxation.

**Methods:** Volunteer participants (N=293) completed a web-based music-listening survey containing 16 questions on Compositional Elements of Relaxation (CER). From the unlabeled audio excerpts isolating and representing variations of each CER isolated from chronologically diverse classical music compositions spanning 400 years, respondents selected the variation perceived to be the most relaxing. Demographics including age, sex, race, education level, occupation, and level of musical training were collected. The 16 CERs identified by music experts included Accentuation, Articulation, Dynamic Range, Familiarity, Interpretive Expertise, Melodic Shape, Meter, Recording Quality, Repetition, Register, Rubato, Tempo, Texture, Timbre, Transition, and Tonality.

**Results:** The web-based music-listening survey was completed by a demographically diverse cohort of 293 volunteer participants. When choosing music with a targeted outcome of relaxation, our investigation identified, evaluated, and validated variations of 16 Compositional Elements of Relaxation that enhanced perceived relaxation. Our data showed that musical compositions with the following intrinsic characteristics promoted relaxation: lack of accentuation, legato articulation, familiarity, pp-mp dynamic range (very soft to medium soft), smooth melodic shape, quadruple meter, high clarity recording, with repetition, middle register, rubato (rhythmic flexibility), medium tempo (80-100 bpm aligning with the human resting heart rate), thin texture, piano or string instrumentation, expert performance, and smooth transition. The most significant factors associated relaxation were legato (connective articulation), an absence of accentuation (strong accents), and rubato (rhythmic flexibility inimitable by computer-generated recordings.) Results from subgroup analysis revealed age, sex, race, education, and musical training differences in preferred music for relaxation. The factors most commonly associated with differences were rubato and texture. Factors that did not differ in any subgroup analysis included Accentuation, Articulation, Interpretive Expertise, Meter, Recording Quality, Repetition, Register, and Timbre. Thin texture was increasingly preferred for relaxation with increasing education and musical training level.

**Conclusions:** Our investigation provided a reproducible theoretical framework for selecting evidence-based qualitative Compositional Elements of Relaxation (CER)—16 parameters isolated and individually assessed as correlated with perceived relaxation. This data-driven music-selecting methodology significantly increases the scientific rigor and the probability of clinical translation for music medicine research with targeted anxiolytic outcomes.

## INTRODUCTION

Music is a powerful psychophysiological modulator of the human stress response, effectively diminishing serum-level stress biomarkers and perceived stress levels by dynamically modulating the hypothalamic-pituitary-adrenal (HPA) axis, autonomic nervous system (ANS) and heart rate variability (HRV). We have previously discovered the physiological mechanism for music anxiolysis in ICU patients (Conrad et al).

A major knowledge gap in the field of music medicine is the dearth of scientifically rigorous reproducible methodology outlining the qualitative musical compositional elements correlated with perceived relaxation, as associated with demographic variables including gender, race, age, preference, level of education, and profession. Using data-driven methodologies and harnessing the expertise of a Grammy-nominated composer, musicologists, and concert artists, we constructed a replicable and precise theoretical framework composed of 16 Compositional Elements of Relaxation (CER), identified as Accentuation, Articulation, Dynamics, Familiarity, Melodic Shape, Meter, Performance Expertise, Recording Quality, Repetition, Register, Rubato, Tempo, Texture, Timbre, Tonality and Transition. 40 excerpts representing 16 CER were evaluated in a web-based music-listening study (N=293).

## METHODS

A panel of musical experts (a Grammy-nominated composer and award-winning concert artist) identified and isolated a set of 16 Compositional Elements of Relaxation (CER)— hypothesized to exert an impact on subjective stress reduction independent of demographic factors including gender, race, age, preference, and level of education. For each of the 16 CERs, our study team composed, prepared and recorded 20-second long excerpts representing 2-4 defined variations of each of the 16 CERs. All musical excerpts used in the study were recorded in Duncan Recital Hall, Rice University in October-December 2020. Participants were recruited via electronic communication that contained a link to an IRB-approved Qualtrics web survey. (PRO#00024163) (https://riceuniversity.co1.qualtrics.com/jfe/form/SV_3qJJO5nKlwOixDv)

This web-based music-listening survey contained 16 questions—each containing 2-4 unlabeled audio excerpts representing variations of each of the CERs. It was distributed via a Houston Methodist Hospital’s Center for Performing Arts Medicine emailing list totaling 5000 people, of which 293 completed the survey (N=293), with a response rate of 5.86%. Inclusion criteria included adults with basic computer literacy over the age of 18. Exclusion criteria included those with deficit, diagnosed cognitive impairment, computer illiteracy, and amusia. From the unlabeled audio excerpts representing variations of each CER, respondents selected the single variation of each CER, which they perceived to be the most relaxing. Demographics including age, sex, race, education level, occupation, and level of musical training were collected.

## RESULTS

The web-based survey was completed by 293 individuals. Of the respondents, 59.7% and 37.6% identified as female and male respectively. White and Asian race comprised 55.8% and 29.8% of the cohort respectively. Predominant occupations were Health Care (18.9%), Performing/Visual Arts (16.2%), and Science/Research (13.2%). Individuals with 10+ years of musical training comprised 30.2% of respondents, whereas 23.6% had no musical training background. For the overall cohort, the most relaxing music included no accentuation (92.7%), legato (93.1%), previously heard pieces (73.2%), pp-mp dynamic (53.5%), master performance (73.6%), smooth melodic shape (69.8%), quadruple meter (41.3%), high clarity recording (86.7%), with repetition (53.6%), middle register (64.1%), with rubato (79.1%), medium tempo (45.2%), thin texture (61.3%), piano/string instrumentation (each 32.6%), and smooth/continuous transitions (59.7%). Minor Tonality was slightly preferred, selected by 50.9% of respondents. (Figure 1) Subgroup analysis (Figures 2 and 3) revealed that by age, differences in relaxation were observed in melodic shape (jagged preference with increasing age, P=0.045) and transition (disjointed preference with increasing age, P=0.014). By sex, differences were observed in dynamic range (f-ff : mp-mf : pp-mp; female 14:63:70, male 11:22:58, P=0.014) and texture (increased preference for thin in males, P=0.035). By White vs Asian race, a difference was observed in rubato (increased with rubato preference in White race, P=0.016). By education, differences were observed in rubato (post-docs prefer with rubato more, P=0.008), texture (thin preference with increasing education, P=0.05), and tonality (P=0.04). By training level, differences were observed in tempo (slower preference in less experienced musicians, P<0.001), texture (thin preference in professionals, P=0.02), and familiarity (new preference with less experience, P=0.01).

**Figure 1:**
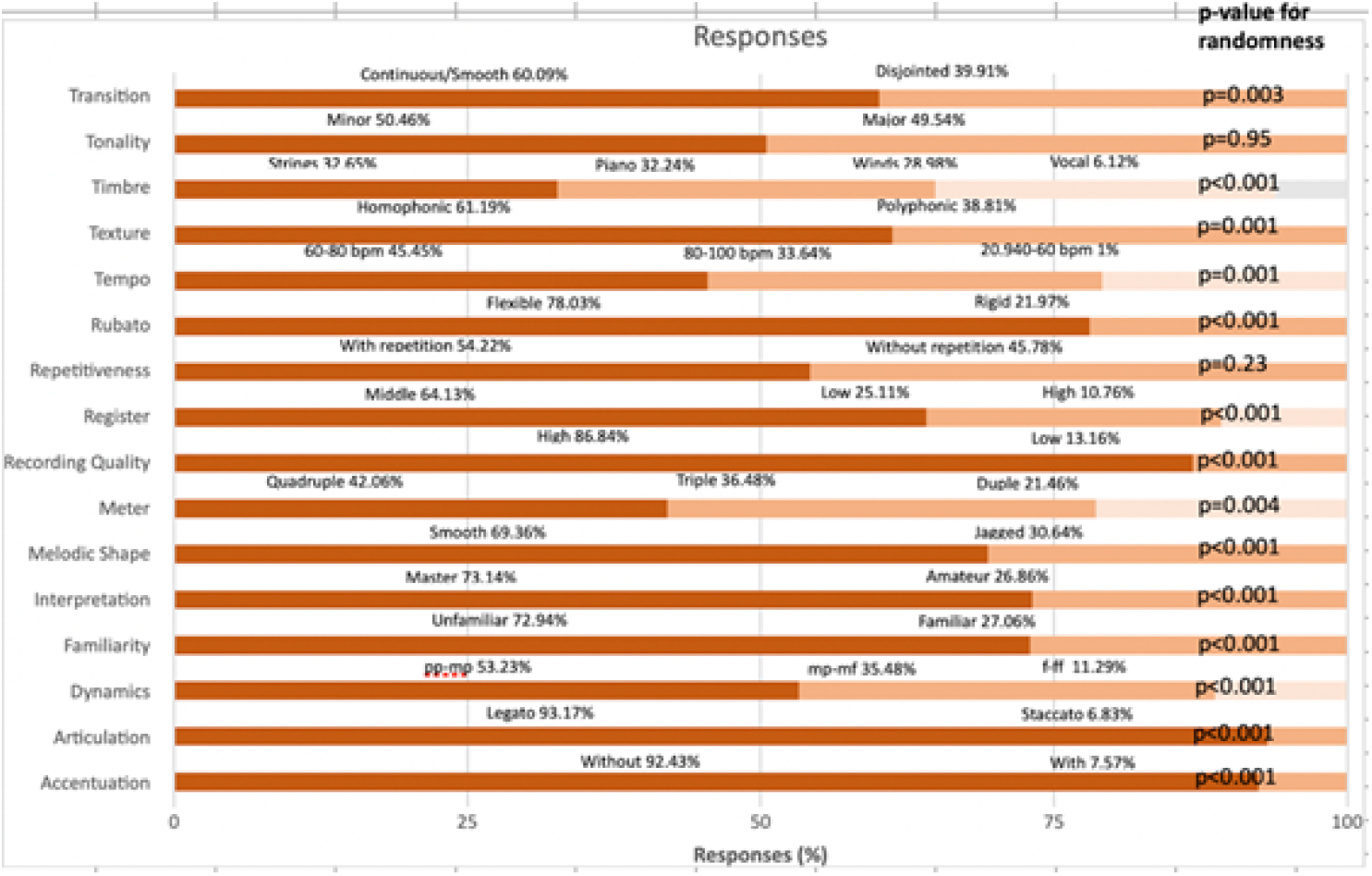
16 Compositional Elements of Relaxation: responses of variations with p-values

**Figure 2:**
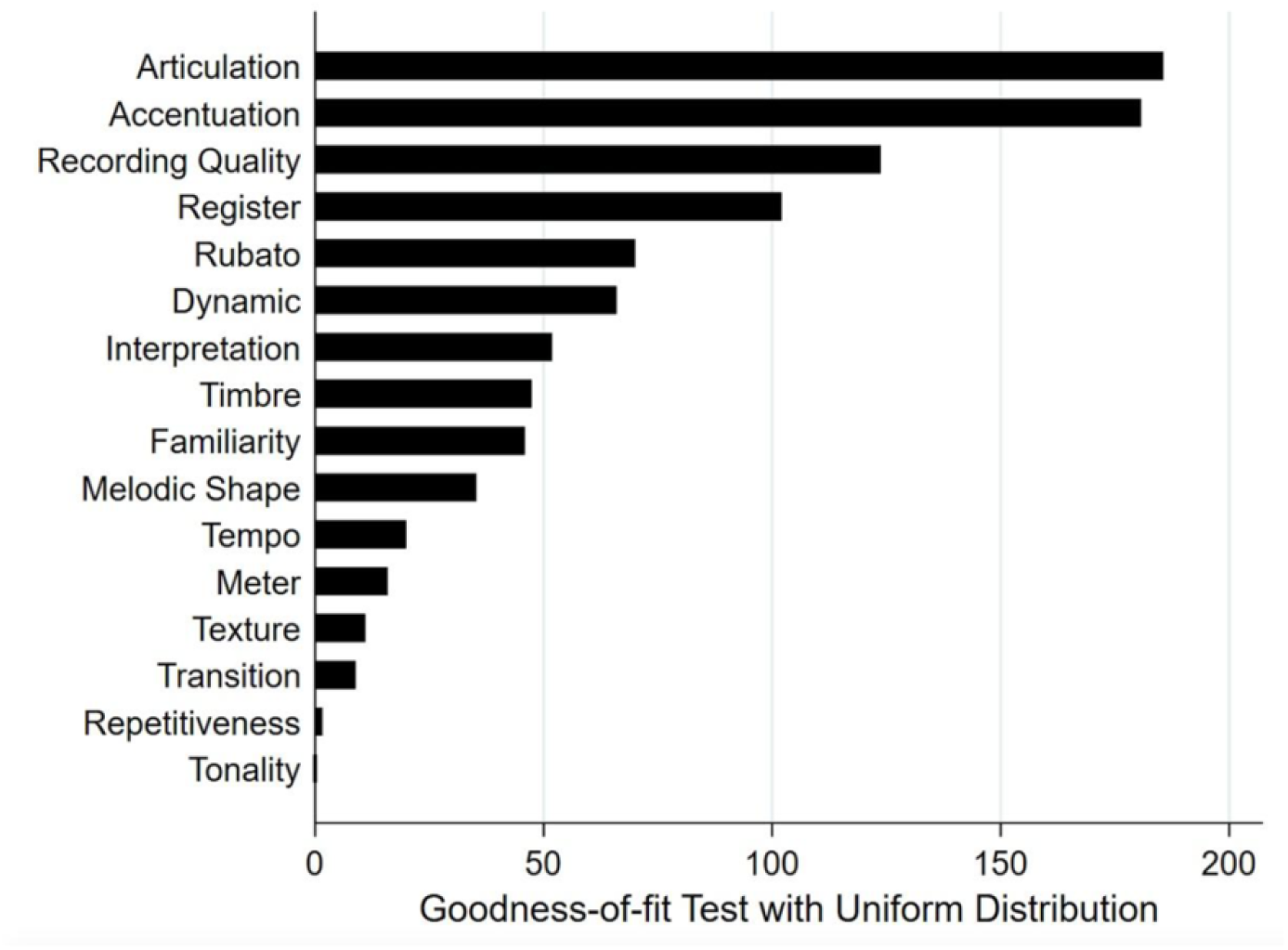
Hierarchical ranking of 16 Compositional Elements of Relaxation

**Figure 3:**
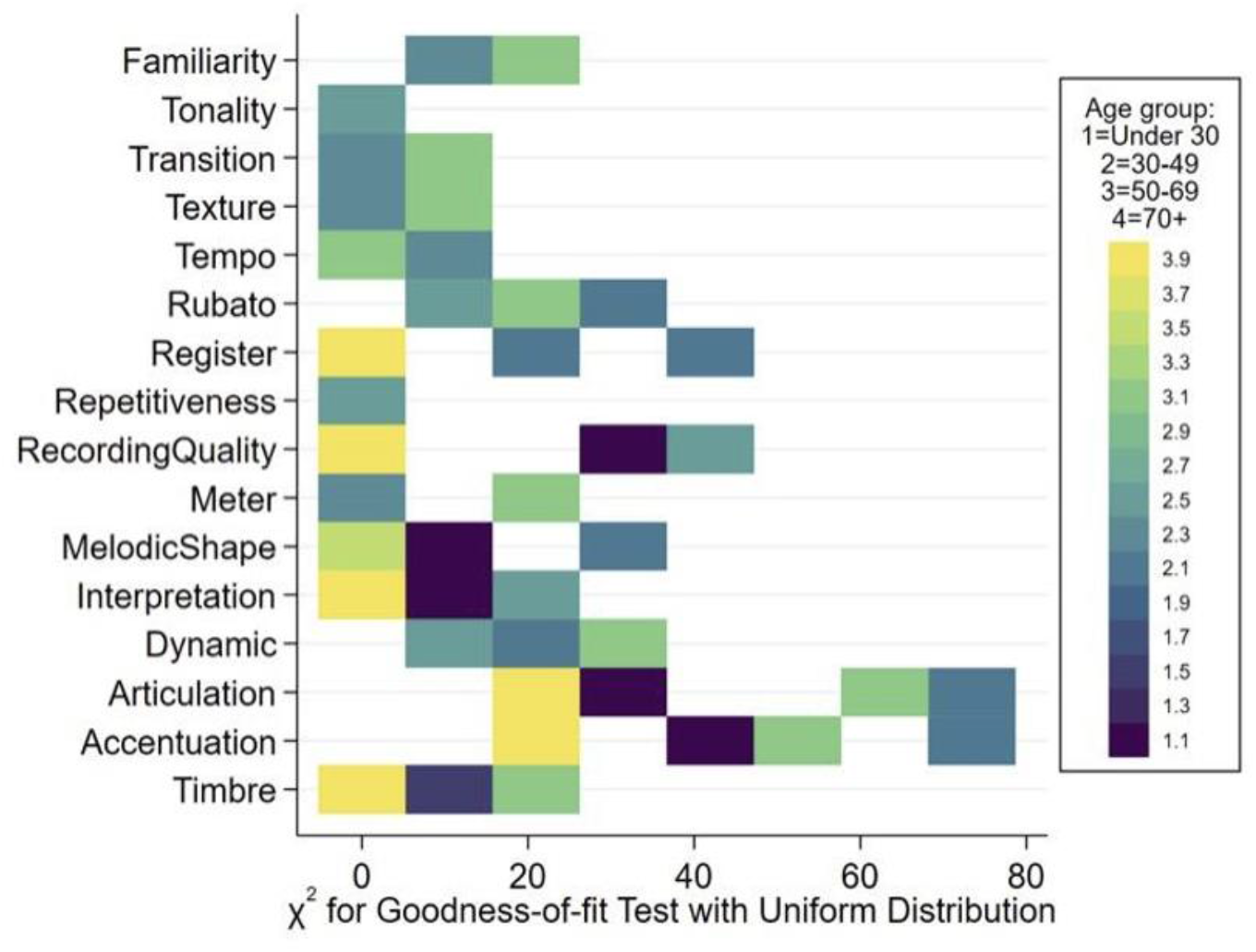
Heat map: association of of 16 Compositional Elements of Relaxation with Age

## DISCUSSION

Our investigation provided evidence that demonstrated, for the first time, precise variations of 16 Compositional Elements of Relaxation (CER) validated to promote perceived relaxation in a demographically diverse cohort. When choosing music with a targeted outcome of relaxation, our data showed that music compositions integrating the following intrinsic characteristics to enhance perceived relaxation: lack of accentuation, legato, previously heard pieces, pp-mp dynamic range (very soft to medium soft), smooth melodic shape, quadruple meter, high clarity recording, with repetition, middle register, rubato (rhythmic flexibility), medium tempo (80-100bpm aligning with the human resting heart rate), thin texture, piano/string instrumentation, expert performance, and smooth transition were associated with relaxing quality of music. The most important and consistent factors associated relaxation were legato (connective articulation), an absence of accentuation (strong accents), and rubato—rhythmic flexibility inimitable by computer-generated recordings.

Results from subgroup analysis revealed age, sex, race, education, and musical training differences in preferred music for relaxation. The factors most commonly associated with differences were rubato and texture. Factors that did not differ in any subgroup analysis included Accentuation, Articulation, Interpretive Expertise, Meter, Recording Quality, Repetition, Register, and Timbre. Thin texture was increasingly preferred for relaxation with increasing education and musical training level.

## LIMITATIONS AND FUTURE DIRECTIONS

This investigation relied on self-reported data based on subjective reaction to and perception of the music excerpts. In the future, a larger subject pool, greater diversity, and a compliance-tracking system would enhance the study’s findings. Another limitation in our investigation is that it does not address the duration or strength of music’s impact on perceived relaxation: are the effects temporary and diminish over time? The length of music exposure may also have an impact on the strength of the results. In this study, music experts from our study group focused on Western classical music, because the majority of prior evidence-based music intervention studies employed this repertoire. In follow-up investigations, it would be worthwhile to account for music preference and direct subjects to choose self-selected music that display similar characteristics as those identified as CER–for instance, smooth jazz and pop songs with a slower tempo. Long-term, we propose to translate our findings into providing a replicable and scientifically rigorous methodology for music repertoire selection in clinical cohorts.

Specifically, we plan to invite expert composers from different musical genres to integrate these validated Compositional Elements of Relaxation into novel compositions, and further test these works in both clinical and non-clinical cohorts. Moreover, we plan to use more quantitative tools to explore and validate our results, such as 7-tesla fMRI imaging, EEG, and pupillometry to evaluate music-induced neurophysiological changes.

## CONCLUSION

Our investigation overcame a major knowledge gap in the field of music therapy and music medicine—a lack of scientifically rigorous, precise, objective, and reproducible theoretical framework outlining specific compositional elements in music that enhance perceived relaxation in a demographically diverse cohort (N=293). In conclusion, this study explored and successfully validated 16 Compositional Elements of Relaxation extracted from a chronologically and culturally diverse selection of music compositions spanning 400 years. Furthermore, our data supported music’s powerful and versatile application as a safe, non-pharmacologic, and cost-effective intervention to enhance relaxation in future music medicine clinical studies.

## Supporting information

CER results table

## Data Availability

All data produced in the present work are contained in the manuscript

https://riceuniversity.co1.qualtrics.com/jfe/form/SV_3qJJO5nKlwOixDv

## Notes

### Competing Interest Statement

The authors have declared no competing interest.

### Funding Statement

This study was funded by Center for Performing Arts Medicine at Houston Methodist Hospital

### Author Declarations

This web based music listening study was approved by the IRB board at Houston Methodist Research Institute. PRO00024163

