## Supplementary material for "Identification and Multivariate Analysis of 16 Compositional Elements of Relaxation—A Web-based Music Listening Study": CER results table

Table 1: Characteristics by education

| | Total | Education | | | p-value |
|---|---|---|---|---|---|
| | | High School | Bechelor | Graduate | |
| | N=240 | N=39 | N=71 | N=130 | |
| Age | | | | | <0.001 |
| Under 30 | 60 (25.00) | 25 (64.10) | 19 (26.76) | 16 (12.31) | |
| 30-49 | 83 (34.58) | 4 (10.26) | 20 (28.17) | 59 (45.38) | |
| 50-69 | 76 (31.67) | 9 (23.08) | 23 (32.39) | 44 (33.85) | |
| 70+ | 21 (8.75) | 1 (2.56) | 9 (12.68) | 11 (8.46) | |
| Gender | | | | | 0.22 |
| Female | 142 (59.17) | 21 (53.85) | 48 (67.61) | 73 (56.15) | |
| Male | 91 (37.92) | 16 (41.03) | 20 (28.17) | 55 (42.31) | |
| Other | 7 (2.92) | 2 (5.13) | 3 (4.23) | 2 (1.54) | |
| Ethnicity | | | | | 0.002 |
| White | 137 (57.08) | 13 (33.33) | 47 (66.20) | 77 (59.23) | |
| African American | 4 (1.67) | 1 (2.56) | 0 (0.00) | 3 (2.31) | |
| Asian | 73 (30.42) | 19 (48.72) | 13 (18.31) | 41 (31.54) | |
| Hispanic | 11 (4.58) | 1 (2.56) | 7 (9.86) | 3 (2.31) | |
| Other | 15 (6.25) | 5 (12.82) | 4 (5.63) | 6 (4.62) | |
| Music-Listening | | | | | 0.87 |
| Never | 20 (8.33) | 3 (7.69) | 7 (9.86) | 10 (7.69) | |
| 1-5 times per week | 117 (48.75) | 16 (41.03) | 37 (52.11) | 64 (49.23) | |
| 6-10 times per week | 43 (17.92) | 7 (17.95) | 12 (16.90) | 24 (18.46) | |
| More than 10 times per week | 60 (25.00) | 13 (33.33) | 15 (21.13) | 32 (24.62) | |
| Musical Training | | | | | 0.030 |
| None | 54 (22.50) | 8 (20.51) | 17 (23.94) | 29 (22.31) | |
| 0-5 years | 68 (28.33) | 11 (28.21) | 24 (33.80) | 33 (25.38) | |
| 10+ years | 38 (15.83) | 8 (20.51) | 11 (15.49) | 19 (14.62) | |
| 6-10 years | 44 (18.33) | 12 (30.77) | 9 (12.68) | 23 (17.69) | |
| Professional musician | 36 (15.00) | 0 (0.00) | 10 (14.08) | 26 (20.00) | |
| Instrument | N=185 | N=30 | N=54 | N=101 | 0.50 |
| Piano | 104 (56.22) | 17 (56.67) | 32 (59.26) | 55 (54.46) | |
| Strings | 24 (12.97) | 7 (23.33) | 8 (14.81) | 9 (8.91) | |
| Vocal | 33 (17.84) | 3 (10.00) | 8 (14.81) | 22 (21.78) | |
| Winds | 18 (9.73) | 2 (6.67) | 4 (7.41) | 12 (11.88) | |
| Percussion | 6 (3.24) | 1 (3.33) | 2 (3.70) | 3 (2.97) | |
| 01_Timbre | N=226 | N=34 | N=69 | N=123 | 0.46 |
| piano | 74 (32.74) | 12 (35.29) | 27 (39.13) | 35 (28.46) | |
| strings | 76 (33.63) | 10 (29.41) | 17 (24.64) | 49 (39.84) | |
| vocal | 15 (6.64) | 3 (8.82) | 4 (5.80) | 8 (6.50) | |
| winds | 61 (26.99) | 9 (26.47) | 21 (30.43) | 31 (25.20) | |
| 02_Accentuation | N=232 | N=36 | N=70 | N=126 | 0.023 |
| with accents | 16 (6.90) | 5 (13.89) | 7 (10.00) | 4 (3.17) | |
| without accents | 216 (93.10) | 31 (86.11) | 63 (90.00) | 122 (96.83) | |
| 03_Articulation | N=231 | N=36 | N=70 | N=125 | 0.40 |
| legato | 216 (93.51) | 32 (88.89) | 67 (95.71) | 117 (93.60) | |

| | | | | | |
|---|---|---|---|---|---|
| staccato | 15 (6.49) | 4 (11.11) | 3 (4.29) | 8 (6.40) | |
| 04_Dynamic | N=230 | N=36 | N=69 | N=125 | 0.48 |
| f-ff | 25 (10.87) | 6 (16.67) | 8 (11.59) | 11 (8.80) | |
| mp-mf | 81 (35.22) | 15 (41.67) | 22 (31.88) | 44 (35.20) | |
| pp-mp | 124 (53.91) | 15 (41.67) | 39 (56.52) | 70 (56.00) | |
| 05_Interpretation | N=224 | N=35 | N=69 | N=120 | 0.39 |
| master performer | 164 (73.21) | 23 (65.71) | 49 (71.01) | 92 (76.67) | |
| mediocre performer | 60 (26.79) | 12 (34.29) | 20 (28.99) | 28 (23.33) | |
| 06_MelodicShape | N=219 | N=35 | N=67 | N=117 | 0.36 |
| jagged | 66 (30.14) | 14 (40.00) | 20 (29.85) | 32 (27.35) | |
| smooth | 153 (69.86) | 21 (60.00) | 47 (70.15) | 85 (72.65) | |
| 07_Meter | N=217 | N=35 | N=65 | N=117 | 0.51 |
| duple | 50 (23.04) | 7 (20.00) | 14 (21.54) | 29 (24.79) | |
| quadruple | 87 (40.09) | 11 (31.43) | 30 (46.15) | 46 (39.32) | |
| triple | 80 (36.87) | 17 (48.57) | 21 (32.31) | 42 (35.90) | |
| 08_RecordingQuality | N=212 | N=33 | N=62 | N=117 | 0.81 |
| high clarity | 184 (86.79) | 29 (87.88) | 55 (88.71) | 100 (85.47) | |
| low clarity | 28 (13.21) | 4 (12.12) | 7 (11.29) | 17 (14.53) | |
| 09_Repetitiveness | N=209 | N=33 | N=62 | N=114 | 0.27 |
| complete repetition | 113 (54.07) | 22 (66.67) | 33 (53.23) | 58 (50.88) | |
| no repetition | 96 (45.93) | 11 (33.33) | 29 (46.77) | 56 (49.12) | |
| 10_Register | N=207 | N=31 | N=62 | N=114 | 0.67 |
| high | 21 (10.14) | 4 (12.90) | 5 (8.06) | 12 (10.53) | |
| low | 53 (25.60) | 9 (29.03) | 19 (30.65) | 25 (21.93) | |
| middle | 133 (64.25) | 18 (58.06) | 38 (61.29) | 77 (67.54) | |
| 11_Rubato | N=207 | N=31 | N=62 | N=114 | 0.049 |
| with rubato | 166 (80.19) | 20 (64.52) | 53 (85.48) | 93 (81.58) | |
| without rubato | 41 (19.81) | 11 (35.48) | 9 (14.52) | 21 (18.42) | |
| 12_Tempo | N=205 | N=31 | N=62 | N=112 | 0.25 |
| fast | 71 (34.63) | 14 (45.16) | 21 (33.87) | 36 (32.14) | |
| medium | 90 (43.90) | 9 (29.03) | 25 (40.32) | 56 (50.00) | |
| slow | 44 (21.46) | 8 (25.81) | 16 (25.81) | 20 (17.86) | |
| 13_Texture | N=205 | N=31 | N=62 | N=112 | 0.15 |
| thick | 82 (40.00) | 15 (48.39) | 29 (46.77) | 38 (33.93) | |
| thin | 123 (60.00) | 16 (51.61) | 33 (53.23) | 74 (66.07) | |
| 14_Transition | N=204 | N=31 | N=62 | N=111 | 0.49 |
| continuous/smooth | 122 (59.80) | 16 (51.61) | 36 (58.06) | 70 (63.06) | |
| disjointed | 82 (40.20) | 15 (48.39) | 26 (41.94) | 41 (36.94) | |
| 15_Tonality | N=204 | N=31 | N=62 | N=111 | 0.96 |
| major | 101 (49.51) | 15 (48.39) | 30 (48.39) | 56 (50.45) | |
| minor | 103 (50.49) | 16 (51.61) | 32 (51.61) | 55 (49.55) | |
| 16_Familiarity | N=204 | N=31 | N=62 | N=111 | 0.98 |
| new excerpt | 51 (25.00) | 8 (25.81) | 15 (24.19) | 28 (25.23) | |
| previously heard excerpt | 153 (75.00) | 23 (74.19) | 47 (75.81) | 83 (74.77) | |

Data are presented as n (%). P-value was obtained from Chi-square test or Fisher’s exact test.

Table 2: Characteristics by age

| | Total | Age | | | | p-value |
|---|---|---|---|---|---|---|
| | | Under 30 | 30-49 | 50-69 | 70+ | |
| | N=240 | N=60 | N=83 | N=76 | N=21 | |
| Gender | | | | | | 0.20 |
| Female | 142 (59.17) | 28 (46.67) | 51 (61.45) | 48 (63.16) | 15 (71.43) | |
| Male | 91 (37.92) | 28 (46.67) | 30 (36.14) | 27 (35.53) | 6 (28.57) | |
| Other | 7 (2.92) | 4 (6.67) | 2 (2.41) | 1 (1.32) | 0 (0.00) | |
| Education | | | | | | <0.001 |
| High School | 39 (16.25) | 25 (41.67) | 4 (4.82) | 9 (11.84) | 1 (4.76) | |
| Bechelor's | 71 (29.58) | 19 (31.67) | 20 (24.10) | 23 (30.26) | 9 (42.86) | |
| Graduate | 130 (54.17) | 16 (26.67) | 59 (71.08) | 44 (57.89) | 11 (52.38) | |
| Ethnicity | | | | | | <0.001 |
| White | 137 (57.08) | 21 (35.00) | 39 (46.99) | 58 (76.32) | 19 (90.48) | |
| African American | 4 (1.67) | 0 (0.00) | 2 (2.41) | 2 (2.63) | 0 (0.00) | |
| Asian | 73 (30.42) | 29 (48.33) | 34 (40.96) | 9 (11.84) | 1 (4.76) | |
| Hispanic | 11 (4.58) | 4 (6.67) | 5 (6.02) | 1 (1.32) | 1 (4.76) | |
| Other | 15 (6.25) | 6 (10.00) | 3 (3.61) | 6 (7.89) | 0 (0.00) | |
| Music Listening | | | | | | 0.10 |
| Never | 20 (8.33) | 5 (8.33) | 3 (3.61) | 8 (10.53) | 4 (19.05) | |
| 1-5 times per week | 117 (48.75) | 23 (38.33) | 51 (61.45) | 33 (43.42) | 10 (47.62) | |
| 6-10 times per week | 43 (17.92) | 11 (18.33) | 13 (15.66) | 16 (21.05) | 3 (14.29) | |
| More than 10 times per week | 60 (25.00) | 21 (35.00) | 16 (19.28) | 19 (25.00) | 4 (19.05) | |
| Music training | | | | | | 0.37 |
| None | 54 (22.50) | 11 (18.33) | 20 (24.10) | 15 (19.74) | 8 (38.10) | |
| 0-5 years | 68 (28.33) | 16 (26.67) | 25 (30.12) | 24 (31.58) | 3 (14.29) | |
| 6-10 years | 44 (18.33) | 14 (23.33) | 14 (16.87) | 11 (14.47) | 5 (23.81) | |
| 10+ years | 38 (15.83) | 12 (20.00) | 8 (9.64) | 14 (18.42) | 4 (19.05) | |
| Professional musician | 36 (15.00) | 7 (11.67) | 16 (19.28) | 12 (15.79) | 1 (4.76) | |
| Instrument | N=185 | N=48 | N=63 | N=61 | N=13 | 0.005 |
| Piano | 104 (56.22) | 32 (66.67) | 29 (46.03) | 36 (59.02) | 7 (53.85) | |
| Strings | 24 (12.97) | 12 (25.00) | 7 (11.11) | 5 (8.20) | 0 (0.00) | |
| Vocal | 33 (17.84) | 2 (4.17) | 16 (25.40) | 13 (21.31) | 2 (15.38) | |
| Winds | 18 (9.73) | 1 (2.08) | 8 (12.70) | 5 (8.20) | 4 (30.77) | |
| Percussion | 6 (3.24) | 1 (2.08) | 3 (4.76) | 2 (3.28) | 0 (0.00) | |
| 01_Timbre | N=226 | N=56 | N=77 | N=72 | N=21 | 0.56 |
| piano | 74 (32.74) | 23 (41.07) | 21 (27.27) | 23 (31.94) | 7 (33.33) | |
| strings | 76 (33.63) | 14 (25.00) | 25 (32.47) | 29 (40.28) | 8 (38.10) | |
| vocal | 15 (6.64) | 4 (7.14) | 6 (7.79) | 5 (6.94) | 0 (0.00) | |
| winds | 61 (26.99) | 15 (26.79) | 25 (32.47) | 15 (20.83) | 6 (28.57) | |
| 02_Accentuation | N=232 | N=59 | N=79 | N=73 | N=21 | 0.31 |
| with accents | 16 (6.90) | 7 (11.86) | 3 (3.80) | 5 (6.85) | 1 (4.76) | |
| without accents | 216 (93.10) | 52 (88.14) | 76 (96.20) | 68 (93.15) | 20 (95.24) | |
| 03_Articulation | N=231 | N=58 | N=79 | N=73 | N=21 | 0.16 |
| legato | 216 (93.51) | 51 (87.93) | 77 (97.47) | 68 (93.15) | 20 (95.24) | |

| | | | | | | |
|---|---|---|---|---|---|---|
| staccato | 15 (6.49) | 7 (12.07) | 2 (2.53) | 5 (6.85) | 1 (4.76) | |
| 04_Dynamic | N=230 | N=58 | N=79 | N=73 | N=20 | 0.31 |
| f-ff | 25 (10.87) | 6 (10.34) | 7 (8.86) | 10 (13.70) | 2 (10.00) | |
| mp-mf | 81 (35.22) | 26 (44.83) | 31 (39.24) | 18 (24.66) | 6 (30.00) | |
| pp-mp | 124 (53.91) | 26 (44.83) | 41 (51.90) | 45 (61.64) | 12 (60.00) | |
| 05_Interpretation | N=224 | N=56 | N=78 | N=70 | N=20 | 0.30 |
| master performer | 164 (73.21) | 38 (67.86) | 60 (76.92) | 54 (77.14) | 12 (60.00) | |
| mediocre performer | 60 (26.79) | 18 (32.14) | 18 (23.08) | 16 (22.86) | 8 (40.00) | |
| 06_MelodicShape | N=219 | N=55 | N=76 | N=69 | N=19 | 0.029 |
| jagged | 66 (30.14) | 15 (27.27) | 15 (19.74) | 28 (40.58) | 8 (42.11) | |
| smooth | 153 (69.86) | 40 (72.73) | 61 (80.26) | 41 (59.42) | 11 (57.89) | |
| 07_Meter | N=217 | N=54 | N=75 | N=69 | N=19 | 0.27 |
| duple | 50 (23.04) | 13 (24.07) | 21 (28.00) | 11 (15.94) | 5 (26.32) | |
| quadruple | 87 (40.09) | 18 (33.33) | 25 (33.33) | 36 (52.17) | 8 (42.11) | |
| triple | 80 (36.87) | 23 (42.59) | 29 (38.67) | 22 (31.88) | 6 (31.58) | |
| 08_RecordingQuality | N=212 | N=51 | N=74 | N=69 | N=18 | 0.36 |
| high clarity | 184 (86.79) | 43 (84.31) | 63 (85.14) | 60 (86.96) | 18 (100.00) | |
| low clarity | 28 (13.21) | 8 (15.69) | 11 (14.86) | 9 (13.04) | 0 (0.00) | |
| 09_Repetitiveness | N=209 | N=51 | N=72 | N=68 | N=18 | 0.12 |
| complete repetition | 113 (54.07) | 31 (60.78) | 32 (44.44) | 42 (61.76) | 8 (44.44) | |
| no repetition | 96 (45.93) | 20 (39.22) | 40 (55.56) | 26 (38.24) | 10 (55.56) | |
| 10_Register | N=207 | N=49 | N=72 | N=68 | N=18 | 0.23 |
| high | 21 (10.14) | 7 (14.29) | 8 (11.11) | 6 (8.82) | 0 (0.00) | |
| low | 53 (25.60) | 10 (20.41) | 14 (19.44) | 24 (35.29) | 5 (27.78) | |
| middle | 133 (64.25) | 32 (65.31) | 50 (69.44) | 38 (55.88) | 13 (72.22) | |
| 11_Rubato | N=207 | N=49 | N=72 | N=68 | N=18 | 0.58 |
| with rubato | 166 (80.19) | 36 (73.47) | 60 (83.33) | 55 (80.88) | 15 (83.33) | |
| without rubato | 41 (19.81) | 13 (26.53) | 12 (16.67) | 13 (19.12) | 3 (16.67) | |
| 12_Tempo | N=205 | N=48 | N=71 | N=68 | N=18 | 0.31 |
| fast | 71 (34.63) | 20 (41.67) | 18 (25.35) | 28 (41.18) | 5 (27.78) | |
| medium | 90 (43.90) | 20 (41.67) | 36 (50.70) | 24 (35.29) | 10 (55.56) | |
| slow | 44 (21.46) | 8 (16.67) | 17 (23.94) | 16 (23.53) | 3 (16.67) | |
| 13_Texture | N=205 | N=48 | N=71 | N=68 | N=18 | 0.56 |
| thick | 82 (40.00) | 18 (37.50) | 28 (39.44) | 26 (38.24) | 10 (55.56) | |
| thin | 123 (60.00) | 30 (62.50) | 43 (60.56) | 42 (61.76) | 8 (44.44) | |
| 14_Transition | N=204 | N=48 | N=71 | N=67 | N=18 | 0.023 |
| continuous/smooth | 122 (59.80) | 28 (58.33) | 40 (56.34) | 48 (71.64) | 6 (33.33) | |
| disjointed | 82 (40.20) | 20 (41.67) | 31 (43.66) | 19 (28.36) | 12 (66.67) | |
| 15_Tonality | N=204 | N=48 | N=71 | N=67 | N=18 | 0.38 |
| major | 101 (49.51) | 26 (54.17) | 39 (54.93) | 29 (43.28) | 7 (38.89) | |
| minor | 103 (50.49) | 22 (45.83) | 32 (45.07) | 38 (56.72) | 11 (61.11) | |
| 16_Familiarity | N=204 | N=48 | N=71 | N=67 | N=18 | 0.28 |
| new excerpt | 51 (25.00) | 12 (25.00) | 23 (32.39) | 13 (19.40) | 3 (16.67) | |
| previously heard excerpt | 153 (75.00) | 36 (75.00) | 48 (67.61) | 54 (80.60) | 15 (83.33) | |

Data are presented as n (%). P-value was obtained from Chi-square test or Fisher's exact test.

Table 3: Characteristics by ethnicity

| | Total | Ethnicity | | | | | p-value |
|---|---|---|---|---|---|---|---|
| | | White | African American | Asian | Hispanic | Other | |
| | N=240 | N=137 | N=4 | N=73 | N=11 | N=15 | |
| Age | | | | | | | <0.001 |
| Under 30 | 60 (25.00) | 21 (15.33) | 0 (0.00) | 29 (39.73) | 4 (36.36) | 6 (40.00) | |
| 30-49 | 83 (34.58) | 39 (28.47) | 2 (50.00) | 34 (46.58) | 5 (45.45) | 3 (20.00) | |
| 50-69 | 76 (31.67) | 58 (42.34) | 2 (50.00) | 9 (12.33) | 1 (9.09) | 6 (40.00) | |
| 70+ | 21 (8.75) | 19 (13.87) | 0 (0.00) | 1 (1.37) | 1 (9.09) | 0 (0.00) | |
| Gender | | | | | | | 0.29 |
| Female | 142 (59.17) | 84 (61.31) | 2 (50.00) | 38 (52.05) | 8 (72.73) | 10 (66.67) | |
| Male | 91 (37.92) | 46 (33.58) | 2 (50.00) | 35 (47.95) | 3 (27.27) | 5 (33.33) | |
| Other | 7 (2.92) | 7 (5.11) | 0 (0.00) | 0 (0.00) | 0 (0.00) | 0 (0.00) | |
| Education | | | | | | | 0.002 |
| High School | 39 (16.25) | 13 (9.49) | 1 (25.00) | 19 (26.03) | 1 (9.09) | 5 (33.33) | |
| Bechelor's | 71 (29.58) | 47 (34.31) | 0 (0.00) | 13 (17.81) | 7 (63.64) | 4 (26.67) | |
| Graduate | 130 (54.17) | 77 (56.20) | 3 (75.00) | 41 (56.16) | 3 (27.27) | 6 (40.00) | |
| Music Listening | | | | | | | 0.61 |
| Never | 20 (8.33) | 14 (10.22) | 0 (0.00) | 6 (8.22) | 0 (0.00) | 0 (0.00) | |
| 1-5 times per week | 117 (48.75) | 66 (48.18) | 3 (75.00) | 37 (50.68) | 4 (36.36) | 7 (46.67) | |
| 6-10 times per week | 43 (17.92) | 27 (19.71) | 0 (0.00) | 10 (13.70) | 4 (36.36) | 2 (13.33) | |
| More than 10 times per week | 60 (25.00) | 30 (21.90) | 1 (25.00) | 20 (27.40) | 3 (27.27) | 6 (40.00) | |
| Music training | | | | | | | 0.18 |
| None | 54 (22.50) | 26 (18.98) | 1 (25.00) | 23 (31.51) | 3 (27.27) | 1 (6.67) | |
| 0-5 years | 68 (28.33) | 36 (26.28) | 1 (25.00) | 22 (30.14) | 5 (45.45) | 4 (26.67) | |
| 6-10 years | 44 (18.33) | 26 (18.98) | 2 (50.00) | 14 (19.18) | 1 (9.09) | 1 (6.67) | |
| 10+ years | 38 (15.83) | 25 (18.25) | 0 (0.00) | 7 (9.59) | 1 (9.09) | 5 (33.33) | |
| Professional musician | 36 (15.00) | 24 (17.52) | 0 (0.00) | 7 (9.59) | 1 (9.09) | 4 (26.67) | |
| Instrument | N=185 | N=111 | N=3 | N=49 | N=8 | N=14 | 0.004 |
| Piano | 104 (56.22) | 65 (58.56) | 1 (33.33) | 31 (63.27) | 4 (50.00) | 3 (21.43) | |
| Strings | 24 (12.97) | 10 (9.01) | 0 (0.00) | 9 (18.37) | 1 (12.50) | 4 (28.57) | |
| Vocal | 33 (17.84) | 20 (18.02) | 0 (0.00) | 6 (12.24) | 1 (12.50) | 6 (42.86) | |
| Winds | 18 (9.73) | 14 (12.61) | 1 (33.33) | 1 (2.04) | 2 (25.00) | 0 (0.00) | |
| Percussion | 6 (3.24) | 2 (1.80) | 1 (33.33) | 2 (4.08) | 0 (0.00) | 1 (7.14) | |
| 01_Timbre | N=226 | N=133 | N=3 | N=67 | N=11 | N=12 | 0.20 |
| piano | 74 (32.74) | 38 (28.57) | 0 (0.00) | 29 (43.28) | 5 (45.45) | 2 (16.67) | |
| strings | 76 (33.63) | 51 (38.35) | 1 (33.33) | 18 (26.87) | 3 (27.27) | 3 (25.00) | |
| vocal | 15 (6.64) | 10 (7.52) | 0 (0.00) | 4 (5.97) | 1 (9.09) | 0 (0.00) | |
| winds | 61 (26.99) | 34 (25.56) | 2 (66.67) | 16 (23.88) | 2 (18.18) | 7 (58.33) | |
| 02_Accentuation | N=232 | N=133 | N=4 | N=70 | N=11 | N=14 | 0.054 |
| with accents | 16 (6.90) | 7 (5.26) | 0 (0.00) | 4 (5.71) | 3 (27.27) | 2 (14.29) | |
| without accents | 216 (93.10) | 126 (94.74) | 4 (100.00) | 66 (94.29) | 8 (72.73) | 12 (85.71) | |
| 03_Articulation | N=231 | N=133 | N=4 | N=70 | N=10 | N=14 | 0.003 |

| | | | | | | | |
|---|---|---|---|---|---|---|---|
| legato | 216 (93.51) | 128 (96.24) | 3 (75.00) | 65 (92.86) | 10 (100.00) | 10 (71.43) | |
| staccato | 15 (6.49) | 5 (3.76) | 1 (25.00) | 5 (7.14) | 0 (0.00) | 4 (28.57) | |
| 04_Dynamic | N=230 | N=132 | N=4 | N=70 | N=10 | N=14 | 0.81 |
| f-ff | 25 (10.87) | 14 (10.61) | 1 (25.00) | 7 (10.00) | 2 (20.00) | 1 (7.14) | |
| mp-mf | 81 (35.22) | 47 (35.61) | 1 (25.00) | 26 (37.14) | 1 (10.00) | 6 (42.86) | |
| pp-mp | 124 (53.91) | 71 (53.79) | 2 (50.00) | 37 (52.86) | 7 (70.00) | 7 (50.00) | |
| 05_Interpretation | N=224 | N=130 | N=3 | N=67 | N=10 | N=14 | 0.59 |
| master performer | 164 (73.21) | 95 (73.08) | 1 (33.33) | 51 (76.12) | 7 (70.00) | 10 (71.43) | |
| mediocre performer | 60 (26.79) | 35 (26.92) | 2 (66.67) | 16 (23.88) | 3 (30.00) | 4 (28.57) | |
| 06_MelodicShape | N=219 | N=127 | N=2 | N=67 | N=10 | N=13 | 0.78 |
| jagged | 66 (30.14) | 40 (31.50) | 1 (50.00) | 18 (26.87) | 2 (20.00) | 5 (38.46) | |
| smooth | 153 (69.86) | 87 (68.50) | 1 (50.00) | 49 (73.13) | 8 (80.00) | 8 (61.54) | |
| 07_Meter | N=217 | N=127 | N=2 | N=67 | N=10 | N=13 | 0.38 |
| duple | 50 (23.04) | 27 (21.26) | 0 (0.00) | 20 (29.85) | 1 (11.11) | 2 (16.67) | |
| quadruple | 87 (40.09) | 57 (44.88) | 0 (0.00) | 22 (32.84) | 4 (44.44) | 4 (33.33) | |
| triple | 80 (36.87) | 43 (33.86) | 2 (100.00) | 25 (37.31) | 4 (44.44) | 6 (50.00) | |
| 08_RecordingQuality | N=212 | N=126 | N=2 | N=66 | N=7 | N=11 | 0.68 |
| high clarity | 184 (86.79) | 111 (88.10) | 2 (100.00) | 56 (84.85) | 5 (71.43) | 10 (90.91) | |
| low clarity | 28 (13.21) | 15 (11.90) | 0 (0.00) | 10 (15.15) | 2 (28.57) | 1 (9.09) | |
| 09_Repetitiveness | N=209 | N=125 | N=2 | N=65 | N=7 | N=10 | 0.52 |
| complete repetition | 113 (54.07) | 64 (51.20) | 1 (50.00) | 36 (55.38) | 4 (57.14) | 8 (80.00) | |
| no repetition | 96 (45.93) | 61 (48.80) | 1 (50.00) | 29 (44.62) | 3 (42.86) | 2 (20.00) | |
| 10_Register | N=207 | N=125 | N=2 | N=63 | N=7 | N=10 | 0.12 |
| high | 21 (10.14) | 9 (7.20) | 0 (0.00) | 9 (14.29) | 0 (0.00) | 3 (30.00) | |
| low | 53 (25.60) | 37 (29.60) | 1 (50.00) | 14 (22.22) | 0 (0.00) | 1 (10.00) | |
| middle | 133 (64.25) | 79 (63.20) | 1 (50.00) | 40 (63.49) | 7 (100.00) | 6 (60.00) | |
| 11_Rubato | N=207 | N=125 | N=2 | N=63 | N=7 | N=10 | 0.12 |
| with rubato | 166 (80.19) | 107 (85.60) | 1 (50.00) | 45 (71.43) | 6 (85.71) | 7 (70.00) | |
| without rubato | 41 (19.81) | 18 (14.40) | 1 (50.00) | 18 (28.57) | 1 (14.29) | 3 (30.00) | |
| 12_Tempo | N=205 | N=124 | N=2 | N=62 | N=7 | N=10 | 0.62 |
| fast | 71 (34.63) | 43 (34.68) | 1 (50.00) | 20 (32.26) | 1 (14.29) | 6 (60.00) | |
| medium | 90 (43.90) | 56 (45.16) | 1 (50.00) | 27 (43.55) | 3 (42.86) | 3 (30.00) | |
| slow | 44 (21.46) | 25 (20.16) | 0 (0.00) | 15 (24.19) | 3 (42.86) | 1 (10.00) | |
| 13_Texture | N=205 | N=124 | N=2 | N=62 | N=7 | N=10 | 0.25 |
| thick | 82 (40.00) | 54 (43.55) | 2 (100.00) | 21 (33.87) | 2 (28.57) | 3 (30.00) | |
| thin | 123 (60.00) | 70 (56.45) | 0 (0.00) | 41 (66.13) | 5 (71.43) | 7 (70.00) | |
| 14_Transition | N=205 | N=124 | N=2 | N=62 | N=7 | N=10 | 0.098 |
| continuous/smooth | 122 (59.80) | 76 (61.79) | 2 (100.00) | 31 (50.00) | 4 (57.14) | 9 (90.00) | |
| disjointed | 82 (40.20) | 47 (38.21) | 0 (0.00) | 31 (50.00) | 3 (42.86) | 1 (10.00) | |
| 15_Tonality | N=205 | N=124 | N=2 | N=62 | N=7 | N=10 | 0.19 |
| major | 101 (49.51) | 61 (49.59) | 0 (0.00) | 34 (54.84) | 1 (14.29) | 5 (50.00) | |
| minor | 103 (50.49) | 62 (50.41) | 2 (100.00) | 28 (45.16) | 6 (85.71) | 5 (50.00) | |
| 16_Familiarity | N=205 | N=124 | N=2 | N=62 | N=7 | N=10 | 0.41 |
| new excerpt | 51 (25.00) | 28 (22.76) | 0 (0.00) | 20 (32.26) | 2 (28.57) | 1 (10.00) | |
| previously heard excerpt | 153 (75.00) | 95 (77.24) | 2 (100.00) | 42 (67.74) | 5 (71.43) | 9 (90.00) | |

Data are presented as n (%). P-value was obtained from Chi-square test or Fisher's exact test.

Table 4: Characteristics by music listening frequency

| | Total | Music listening | | | | p-value |
|---|---|---|---|---|---|---|
| | | Never | 1-5 times per week | 6-10 times per week | More than 10 times per week | |
| | **N=240** | **N=20** | **N=117** | **N=43** | **N=60** | |
| Age | | | | | | 0.10 |
| Under 30 | 60 (25.00) | 5 (25.00) | 23 (19.66) | 11 (25.58) | 21 (35.00) | |
| 30-49 | 83 (34.58) | 3 (15.00) | 51 (43.59) | 13 (30.23) | 16 (26.67) | |
| 50-69 | 76 (31.67) | 8 (40.00) | 33 (28.21) | 16 (37.21) | 19 (31.67) | |
| 70+ | 21 (8.75) | 4 (20.00) | 10 (8.55) | 3 (6.98) | 4 (6.67) | |
| Gender | | | | | | 0.91 |
| Female | 142 (59.17) | 13 (65.00) | 71 (60.68) | 25 (58.14) | 33 (55.00) | |
| Male | 91 (37.92) | 7 (35.00) | 43 (36.75) | 17 (39.53) | 24 (40.00) | |
| Other | 7 (2.92) | 0 (0.00) | 3 (2.56) | 1 (2.33) | 3 (5.00) | |
| Ethnicity | | | | | | 0.61 |
| White | 137 (57.08) | 14 (70.00) | 66 (56.41) | 27 (62.79) | 30 (50.00) | |
| African American | 4 (1.67) | 0 (0.00) | 3 (2.56) | 0 (0.00) | 1 (1.67) | |
| Asian | 73 (30.42) | 6 (30.00) | 37 (31.62) | 10 (23.26) | 20 (33.33) | |
| Hispanic | 11 (4.58) | 0 (0.00) | 4 (3.42) | 4 (9.30) | 3 (5.00) | |
| Other | 15 (6.25) | 0 (0.00) | 7 (5.98) | 2 (4.65) | 6 (10.00) | |
| Education | | | | | | 0.87 |
| High School | 39 (16.25) | 3 (15.00) | 16 (13.68) | 7 (16.28) | 13 (21.67) | |
| Bechelor's | 71 (29.58) | 7 (35.00) | 37 (31.62) | 12 (27.91) | 15 (25.00) | |
| Graduate | 130 (54.17) | 10 (50.00) | 64 (54.70) | 24 (55.81) | 32 (53.33) | |
| MusicalTraining | | | | | | 0.021 |
| 0-5 years | 68 (28.33) | 3 (15.00) | 33 (28.21) | 16 (37.21) | 16 (26.67) | |
| 10+ years | 38 (15.83) | 1 (5.00) | 13 (11.11) | 11 (25.58) | 13 (21.67) | |
| 6-10 years | 44 (18.33) | 3 (15.00) | 23 (19.66) | 6 (13.95) | 12 (20.00) | |
| None | 54 (22.50) | 9 (45.00) | 33 (28.21) | 5 (11.63) | 7 (11.67) | |
| Professional musician | 36 (15.00) | 4 (20.00) | 15 (12.82) | 5 (11.63) | 12 (20.00) | |
| Instrument | N=185 | N=11 | N=84 | N=38 | N=52 | 0.77 |
| Piano | 104 (56.22) | 5 (45.45) | 45 (53.57) | 22 (57.89) | 32 (61.54) | |
| Strings | 24 (12.97) | 2 (18.18) | 11 (13.10) | 3 (7.89) | 8 (15.38) | |
| Vocal | 33 (17.84) | 2 (18.18) | 17 (20.24) | 8 (21.05) | 6 (11.54) | |
| Winds | 18 (9.73) | 2 (18.18) | 8 (9.52) | 5 (13.16) | 3 (5.77) | |
| Percussion | 6 (3.24) | 0 (0.00) | 3 (3.57) | 0 (0.00) | 3 (5.77) | |
| 01_Timbre | N=226 | N=20 | N=111 | N=41 | N=54 | 0.11 |
| piano | 74 (32.74) | 5 (25.00) | 35 (31.53) | 12 (29.27) | 22 (40.74) | |
| strings | 76 (33.63) | 12 (60.00) | 35 (31.53) | 11 (26.83) | 18 (33.33) | |
| vocal | 15 (6.64) | 0 (0.00) | 8 (7.21) | 2 (4.88) | 5 (9.26) | |
| winds | 61 (26.99) | 3 (15.00) | 33 (29.73) | 16 (39.02) | 9 (16.67) | |
| 02_Accentuation | N=232 | N=20 | N=114 | N=42 | N=56 | 0.56 |
| with accents | 16 (6.90) | 1 (5.00) | 7 (6.14) | 5 (11.90) | 3 (5.36) | |

| | | | | | | |
|---|---|---|---|---|---|---|
| without accents | 216 (93.10) | 19 (95.00) | 107 (93.86) | 37 (88.10) | 53 (94.64) | |
| 03_Articulation | N=231 | N=20 | N=114 | N=41 | N=56 | 0.52 |
| legato | 216 (93.51) | 19 (95.00) | 104 (91.23) | 40 (97.56) | 53 (94.64) | |
| staccato | 15 (6.49) | 1 (5.00) | 10 (8.77) | 1 (2.44) | 3 (5.36) | |
| 04_Dynamic | N=230 | N=20 | N=113 | N=41 | N=56 | 0.12 |
| f-ff | 25 (10.87) | 3 (15.00) | 11 (9.73) | 8 (19.51) | 3 (5.36) | |
| mp-mf | 81 (35.22) | 3 (15.00) | 39 (34.51) | 14 (34.15) | 25 (44.64) | |
| pp-mp | 124 (53.91) | 14 (70.00) | 63 (55.75) | 19 (46.34) | 28 (50.00) | |
| 05_Interpretation | N=224 | N=20 | N=111 | N=39 | N=54 | 0.30 |
| master performer | 164 (73.21) | 12 (60.00) | 86 (77.48) | 26 (66.67) | 40 (74.07) | |
| mediocre performer | 60 (26.79) | 8 (40.00) | 25 (22.52) | 13 (33.33) | 14 (25.93) | |
| 06_MelodicShape | N=219 | N=19 | N=108 | N=39 | N=53 | 0.86 |
| jagged | 66 (30.14) | 6 (31.58) | 32 (29.63) | 10 (25.64) | 18 (33.96) | |
| smooth | 153 (69.86) | 13 (68.42) | 76 (70.37) | 29 (74.36) | 35 (66.04) | |
| 07_Meter | N=217 | N=19 | N=106 | N=39 | N=53 | 0.36 |
| duple | 50 (23.04) | 1 (5.26) | 24 (22.64) | 11 (28.21) | 14 (26.42) | |
| quadruple | 87 (40.09) | 12 (63.16) | 42 (39.62) | 13 (33.33) | 20 (37.74) | |
| triple | 80 (36.87) | 6 (31.58) | 40 (37.74) | 15 (38.46) | 19 (35.85) | |
| 08_RecordingQuality | N=212 | N=19 | N=102 | N=39 | N=52 | 0.68 |
| high clarity | 184 (86.79) | 15 (78.95) | 90 (88.24) | 33 (84.62) | 46 (88.46) | |
| low clarity | 28 (13.21) | 4 (21.05) | 12 (11.76) | 6 (15.38) | 6 (11.54) | |
| 09_Repetitiveness | N=209 | N=19 | N=100 | N=39 | N=51 | 0.48 |
| complete repetition | 113 (54.07) | 11 (57.89) | 51 (51.00) | 19 (48.72) | 32 (62.75) | |
| no repetition | 96 (45.93) | 8 (42.11) | 49 (49.00) | 20 (51.28) | 19 (37.25) | |
| 10_Register | N=207 | N=19 | N=99 | N=39 | N=50 | 0.59 |
| high | 21 (10.14) | 1 (5.26) | 10 (10.10) | 3 (7.69) | 7 (14.00) | |
| low | 53 (25.60) | 8 (42.11) | 25 (25.25) | 8 (20.51) | 12 (24.00) | |
| middle | 133 (64.25) | 10 (52.63) | 64 (64.65) | 28 (71.79) | 31 (62.00) | |
| 11_Rubato | N=207 | N=19 | N=99 | N=39 | N=50 | 0.56 |
| with rubato | 166 (80.19) | 15 (78.95) | 83 (83.84) | 31 (79.49) | 37 (74.00) | |
| without rubato | 41 (19.81) | 4 (21.05) | 16 (16.16) | 8 (20.51) | 13 (26.00) | |
| 12_Tempo | N=205 | N=18 | N=99 | N=39 | N=49 | 0.054 |
| fast | 71 (34.63) | 4 (22.22) | 26 (26.26) | 20 (51.28) | 21 (42.86) | |
| medium | 90 (43.90) | 11 (61.11) | 46 (46.46) | 13 (33.33) | 20 (40.82) | |
| slow | 44 (21.46) | 3 (16.67) | 27 (27.27) | 6 (15.38) | 8 (16.33) | |
| 13_Texture | | | | | | 0.032 |
| thick | 82 (40.00) | 8 (44.44) | 49 (49.49) | 11 (28.21) | 14 (28.57) | |
| thin | 123 (60.00) | 10 (55.56) | 50 (50.51) | 28 (71.79) | 35 (71.43) | |
| 14_Transition | N=205 | N=18 | N=99 | N=39 | N=49 | 0.21 |
| continuous/smooth | 122 (59.80) | 11 (61.11) | 52 (52.53) | 26 (68.42) | 33 (67.35) | |
| disjointed | 82 (40.20) | 7 (38.89) | 47 (47.47) | 12 (31.58) | 16 (32.65) | |
| 15_Tonality | N=205 | N=18 | N=99 | N=39 | N=49 | 0.23 |
| major | 101 (49.51) | 10 (55.56) | 52 (52.53) | 21 (55.26) | 18 (36.73) | |
| minor | 103 (50.49) | 8 (44.44) | 47 (47.47) | 17 (44.74) | 31 (63.27) | |
| 16_Familiarity | N=205 | N=18 | N=99 | N=39 | N=49 | 0.26 |
| new excerpt | 51 (25.00) | 7 (38.89) | 27 (27.27) | 6 (15.79) | 11 (22.45) | |

|  |  |  |  |  |  |
|---|---|---|---|---|---|
| previously heard excerpt | 153 (75.00) | 11 (61.11) | 72 (72.73) | 32 (84.21) | 38 (77.55) |

Data are presented as n (%). P-value was obtained from Chi-square test or Fisher's exact test.

Table 5: Characteristics by music training

|  | Total | Music training |  |  |  |  | p-value |
|---|---|---|---|---|---|---|---|
|  |  | None | 0-5 years | 6-10 years | 10+ years | Professional musician |  |
|  | N=240 | N=54 | N=68 | N=44 | N=38 | N=36 |  |
| Age |  |  |  |  |  |  | 0.37 |
| Under 30 | 60 (25.00) | 11 (20.37) | 16 (23.53) | 14 (31.82) | 12 (31.58) | 7 (19.44) |  |
| 30-49 | 83 (34.58) | 20 (37.04) | 25 (36.76) | 14 (31.82) | 8 (21.05) | 16 (44.44) |  |
| 50-69 | 76 (31.67) | 15 (27.78) | 24 (35.29) | 11 (25.00) | 14 (36.84) | 12 (33.33) |  |
| 70+ | 21 (8.75) | 8 (14.81) | 3 (4.41) | 5 (11.36) | 4 (10.53) | 1 (2.78) |  |
| Gender |  |  |  |  |  |  | 0.41 |
| Female | 142 (59.17) | 33 (61.11) | 40 (58.82) | 29 (65.91) | 25 (65.79) | 15 (41.67) |  |
| Male | 91 (37.92) | 20 (37.04) | 27 (39.71) | 14 (31.82) | 11 (28.95) | 19 (52.78) |  |
| Other | 7 (2.92) | 1 (1.85) | 1 (1.47) | 1 (2.27) | 2 (5.26) | 2 (5.56) |  |
| Ethnicity |  |  |  |  |  |  | 0.18 |
| White | 137 (57.08) | 26 (48.15) | 36 (52.94) | 26 (59.09) | 25 (65.79) | 24 (66.67) |  |
| African American | 4 (1.67) | 1 (1.85) | 1 (1.47) | 2 (4.55) | 0 (0.00) | 0 (0.00) |  |
| Asian | 73 (30.42) | 23 (42.59) | 22 (32.35) | 14 (31.82) | 7 (18.42) | 7 (19.44) |  |
| Hispanic | 11 (4.58) | 3 (5.56) | 5 (7.35) | 1 (2.27) | 1 (2.63) | 1 (2.78) |  |
| Other | 15 (6.25) | 1 (1.85) | 4 (5.88) | 1 (2.27) | 5 (13.16) | 4 (11.11) |  |
| Education |  |  |  |  |  |  | 0.068 |
| High School | 39 (16.25) | 8 (14.81) | 11 (16.18) | 12 (27.27) | 8 (21.05) | 0 (0.00) |  |
| Bechelor's | 71 (29.58) | 17 (31.48) | 24 (35.29) | 9 (20.45) | 11 (28.95) | 10 (27.78) |  |
| Graduate | 130 (54.17) | 29 (53.70) | 33 (48.53) | 23 (52.27) | 19 (50.00) | 26 (72.22) |  |
| Music Listening |  |  |  |  |  |  | 0.021 |
| Never | 20 (8.33) | 9 (16.67) | 3 (4.41) | 3 (6.82) | 1 (2.63) | 4 (11.11) |  |
| 1-5 times per week | 117 (48.75) | 33 (61.11) | 33 (48.53) | 23 (52.27) | 13 (34.21) | 15 (41.67) |  |
| 6-10 times per week | 43 (17.92) | 5 (9.26) | 16 (23.53) | 6 (13.64) | 11 (28.95) | 5 (13.89) |  |
| More than 10 times per week | 60 (25.00) | 7 (12.96) | 16 (23.53) | 12 (27.27) | 13 (34.21) | 12 (33.33) |  |
| Instrument | N=185 | N=0 | N=67 | N=44 | N=38 | N=36 | 0.83 |
| Piano | 104 (56.22) |  | 33 (49.25) | 26 (59.09) | 24 (63.16) | 21 (58.33) |  |
| Strings | 24 (12.97) |  | 9 (13.43) | 7 (15.91) | 4 (10.53) | 4 (11.11) |  |
| Vocal | 33 (17.84) |  | 16 (23.88) | 5 (11.36) | 6 (15.79) | 6 (16.67) |  |
| Winds | 18 (9.73) |  | 8 (11.94) | 3 (6.82) | 3 (7.89) | 4 (11.11) |  |
| Percussion | 6 (3.24) |  | 1 (1.49) | 3 (6.82) | 1 (2.63) | 1 (2.78) |  |
| 01_Timbre | N=226 | N=52 | N=65 | N=41 | N=36 | N=32 | 0.16 |

| | | | | | | | |
|---|---|---|---|---|---|---|---|
| piano | 74 (32.74) | 19 (36.54) | 22 (33.85) | 10 (24.39) | 12 (33.33) | 11 (34.38) | |
| strings | 76 (33.63) | 21 (40.38) | 22 (33.85) | 16 (39.02) | 10 (27.78) | 7 (21.88) | |
| vocal | 15 (6.64) | 3 (5.77) | 6 (9.23) | 1 (2.44) | 0 (0.00) | 5 (15.63) | |
| winds | 61 (26.99) | 9 (17.31) | 15 (23.08) | 14 (34.15) | 14 (38.89) | 9 (28.13) | |
| 02_Accentuation | N=232 | N=53 | N=64 | N=43 | N=37 | N=35 | 0.22 |
| with accents | 16 (6.90) | 5 (9.43) | 7 (10.94) | 3 (6.98) | 1 (2.70) | 0 (0.00) | |
| without accents | 216 (93.10) | 48 (90.57) | 57 (89.06) | 40 (93.02) | 36 (97.30) | 35 (100.00) | |
| 03_Articulation | N=231 | N=53 | N=63 | N=43 | N=37 | N=35 | 0.80 |
| legato | 216 (93.51) | 49 (92.45) | 61 (96.83) | 40 (93.02) | 34 (91.89) | 32 (91.43) | |
| staccato | 15 (6.49) | 4 (7.55) | 2 (3.17) | 3 (6.98) | 3 (8.11) | 3 (8.57) | |
| 04_Dynamic | N=230 | N=53 | N=63 | N=43 | N=37 | N=35 | 0.55 |
| f-ff | 25 (10.87) | 6 (11.32) | 7 (11.11) | 4 (9.52) | 5 (13.51) | 3 (8.57) | |
| mp-mf | 81 (35.22) | 16 (30.19) | 18 (28.57) | 14 (33.33) | 18 (48.65) | 15 (42.86) | |
| pp-mp | 124 (53.91) | 31 (58.49) | 38 (60.32) | 24 (57.14) | 14 (37.84) | 17 (48.57) | |
| 05_Interpretation | N=224 | N=51 | N=61 | N=41 | N=37 | N=34 | 0.046 |
| master performer | 164 (73.21) | 34 (66.67) | 39 (63.93) | 36 (87.80) | 27 (72.97) | 28 (82.35) | |
| mediocre performer | 60 (26.79) | 17 (33.33) | 22 (36.07) | 5 (12.20) | 10 (27.03) | 6 (17.65) | |
| 06_MelodicShape | N=219 | N=49 | N=60 | N=40 | N=37 | N=33 | 0.62 |
| jagged | 66 (30.14) | 17 (34.69) | 16 (26.67) | 14 (35.00) | 12 (32.43) | 7 (21.21) | |
| smooth | 153 (69.86) | 32 (65.31) | 44 (73.33) | 26 (65.00) | 25 (67.57) | 26 (78.79) | |
| 07_Meter | N=217 | N=47 | N=60 | N=40 | N=37 | N=33 | 0.33 |
| duple | 50 (23.04) | 12 (25.53) | 13 (21.67) | 8 (20.00) | 11 (29.73) | 6 (18.18) | |
| quadruple | 87 (40.09) | 20 (42.55) | 30 (50.00) | 17 (42.50) | 9 (24.32) | 11 (33.33) | |
| triple | 80 (36.87) | 15 (31.91) | 17 (28.33) | 15 (37.50) | 17 (45.95) | 16 (48.48) | |
| 08_RecordingQuality | N=212 | N=46 | N=59 | N=39 | N=35 | N=33 | 0.052 |
| high clarity | 184 (86.79) | 41 (89.13) | 47 (79.66) | 31 (79.49) | 34 (97.14) | 31 (93.94) | |
| low clarity | 28 (13.21) | 5 (10.87) | 12 (20.34) | 8 (20.51) | 1 (2.86) | 2 (6.06) | |
| 09_Repetitiveness | N=209 | N=46 | N=59 | N=36 | N=35 | N=33 | 0.69 |
| complete repetition | 113 (54.07) | 26 (56.52) | 33 (55.93) | 21 (58.33) | 19 (54.29) | 14 (42.42) | |
| no repetition | 96 (45.93) | 20 (43.48) | 26 (44.07) | 15 (41.67) | 16 (45.71) | 19 (57.58) | |
| 10_Register | N=207 | N=46 | N=58 | N=35 | N=35 | N=33 | 0.46 |
| high | 21 (10.14) | 2 (4.35) | 6 (10.34) | 4 (11.43) | 4 (11.43) | 5 (15.15) | |
| low | 53 (25.60) | 12 (26.09) | 14 (24.14) | 11 (31.43) | 12 (34.29) | 4 (12.12) | |
| middle | 133 (64.25) | 32 (69.57) | 38 (65.52) | 20 (57.14) | 19 (54.29) | 24 (72.73) | |
| 11_Rubato | N=207 | N=46 | N=58 | N=35 | N=35 | N=33 | 0.43 |
| with rubato | 166 (80.19) | 37 (80.43) | 45 (77.59) | 30 (85.71) | 25 (71.43) | 29 (87.88) | |
| without rubato | 41 (19.81) | 9 (19.57) | 13 (22.41) | 5 (14.29) | 10 (28.57) | 4 (12.12) | |
| 12_Tempo | N=205 | N=46 | N=57 | N=35 | N=35 | N=32 | 0.004 |
| fast | 71 (34.63) | 15 (32.61) | 11 (19.30) | 15 (42.86) | 18 (51.43) | 12 (37.50) | |
| medium | 90 (43.90) | 21 (45.65) | 24 (42.11) | 13 (37.14) | 15 (42.86) | 17 (53.13) | |
| slow | 44 (21.46) | 10 (21.74) | 22 (38.60) | 7 (20.00) | 2 (5.71) | 3 (9.38) | |
| 13_Texture | N=205 | N=46 | N=57 | N=35 | N=35 | N=32 | 0.014 |
| thick | 82 (40.00) | 20 (43.48) | 28 (49.12) | 15 (42.86) | 15 (42.86) | 4 (12.50) | |
| thin | 123 (60.00) | 26 (56.52) | 29 (50.88) | 20 (57.14) | 20 (57.14) | 28 (87.50) | |

| | | | | | | | |
|---|---|---|---|---|---|---|---|
| 14_Transition | N=204 | N=46 | N=57 | N=35 | N=35 | N=31 | 0.10 |
| continuous/smooth | 122 (59.80) | 26 (56.52) | 37 (64.91) | 15 (42.86) | 21 (60.00) | 23 (74.19) | |
| disjointed | 82 (40.20) | 20 (43.48) | 20 (35.09) | 20 (57.14) | 14 (40.00) | 8 (25.81) | |
| 15_Tonality | N=204 | N=46 | N=57 | N=35 | N=35 | N=31 | 0.83 |
| major | 101 (49.51) | 25 (54.35) | 25 (43.86) | 19 (54.29) | 17 (48.57) | 15 (48.39) | |
| minor | 103 (50.49) | 21 (45.65) | 32 (56.14) | 16 (45.71) | 18 (51.43) | 16 (51.61) | |
| 16_Familiarity | N=204 | N=46 | N=57 | N=35 | N=35 | N=31 | 0.022 |
| new excerpt | 51 (25.00) | 14 (30.43) | 21 (36.84) | 8 (22.86) | 3 (8.57) | 5 (16.13) | |
| previously heard excerpt | 153 (75.00) | 32 (69.57) | 36 (63.16) | 27 (77.14) | 32 (91.43) | 26 (83.87) | |

Data are presented as n (%). P-value was obtained from Chi-square test or Fisher's exact test.

Table 6: Characteristics by music instrument played

| | Total | Instrument | | | | | p-value |
|---|---|---|---|---|---|---|---|
| | | Piano | Strings | Vocal | Winds | Percussion | |
| | N=185 | N=104 | N=24 | N=33 | N=18 | N=6 | |
| Age | | | | | | | 0.005 |
| Under 30 | 48 (25.95) | 32 (30.77) | 12 (50.00) | 2 (6.06) | 1 (5.56) | 1 (16.67) | |
| 30-49 | 63 (34.05) | 29 (27.88) | 7 (29.17) | 16 (48.48) | 8 (44.44) | 3 (50.00) | |
| 50-69 | 61 (32.97) | 36 (34.62) | 5 (20.83) | 13 (39.39) | 5 (27.78) | 2 (33.33) | |
| 70+ | 13 (7.03) | 7 (6.73) | 0 (0.00) | 2 (6.06) | 4 (22.22) | 0 (0.00) | |
| Gender | | | | | | | 0.53 |
| Female | 109 (58.92) | 65 (62.50) | 12 (50.00) | 22 (66.67) | 8 (44.44) | 2 (33.33) | |
| Male | 70 (37.84) | 35 (33.65) | 11 (45.83) | 11 (33.33) | 9 (50.00) | 4 (66.67) | |
| Other | 6 (3.24) | 4 (3.85) | 1 (4.17) | 0 (0.00) | 1 (5.56) | 0 (0.00) | |
| Ethnicity | | | | | | | 0.004 |
| White | 111 (60.00) | 65 (62.50) | 10 (41.67) | 20 (60.61) | 14 (77.78) | 2 (33.33) | |
| African American | 3 (1.62) | 1 (0.96) | 0 (0.00) | 0 (0.00) | 1 (5.56) | 1 (16.67) | |
| Asian | 49 (26.49) | 31 (29.81) | 9 (37.50) | 6 (18.18) | 1 (5.56) | 2 (33.33) | |
| Hispanic | 8 (4.32) | 4 (3.85) | 1 (4.17) | 1 (3.03) | 2 (11.11) | 0 (0.00) | |
| Other | 14 (7.57) | 3 (2.88) | 4 (16.67) | 6 (18.18) | 0 (0.00) | 1 (16.67) | |
| Education | | | | | | | 0.50 |
| High School | 30 (16.22) | 17 (16.35) | 7 (29.17) | 3 (9.09) | 2 (11.11) | 1 (16.67) | |
| Bechelor's | 54 (29.19) | 32 (30.77) | 8 (33.33) | 8 (24.24) | 4 (22.22) | 2 (33.33) | |
| Graduate | 101 (54.59) | 55 (52.88) | 9 (37.50) | 22 (66.67) | 12 (66.67) | 3 (50.00) | |
| Music Listening | | | | | | | 0.77 |
| Never | 11 (5.95) | 5 (4.81) | 2 (8.33) | 2 (6.06) | 2 (11.11) | 0 (0.00) | |
| 1-5 times per week | 84 (45.41) | 45 (43.27) | 11 (45.83) | 17 (51.52) | 8 (44.44) | 3 (50.00) | |
| 6-10 times per week | 38 (20.54) | 22 (21.15) | 3 (12.50) | 8 (24.24) | 5 (27.78) | 0 (0.00) | |
| More than 10 times per week | 52 (28.11) | 32 (30.77) | 8 (33.33) | 6 (18.18) | 3 (16.67) | 3 (50.00) | |
| Music training | | | | | | | 0.83 |
| 0-5 years | 67 (36.22) | 33 (31.73) | 9 (37.50) | 16 (48.48) | 8 (44.44) | 1 (16.67) | |
| 6-10 years | 44 (23.78) | 26 (25.00) | 7 (29.17) | 5 (15.15) | 3 (16.67) | 3 (50.00) | |

| | | | | | | | |
|---|---|---|---|---|---|---|---|
| 10+ years | 38 (20.54) | 24 (23.08) | 4 (16.67) | 6 (18.18) | 3 (16.67) | 1 (16.67) | |
| Professional musician | 36 (19.46) | 21 (20.19) | 4 (16.67) | 6 (18.18) | 4 (22.22) | 1 (16.67) | |
| 01_Timbre | N=174 | N=98 | N=20 | N=32 | N=18 | N=6 | 0.12 |
| piano | 55 (31.61) | 37 (37.76) | 6 (30.00) | 6 (18.75) | 6 (33.33) | 0 (0.00) | |
| strings | 55 (31.61) | 27 (27.55) | 6 (30.00) | 17 (53.13) | 2 (11.11) | 3 (50.00) | |
| vocal | 12 (6.90) | 7 (7.14) | 2 (10.00) | 1 (3.13) | 1 (5.56) | 1 (16.67) | |
| winds | 52 (29.89) | 27 (27.55) | 6 (30.00) | 8 (25.00) | 9 (50.00) | 2 (33.33) | |
| 02_Accentuation | N=179 | N=100 | N=23 | N=32 | N=18 | N=6 | 0.008 |
| with accents | 11 (6.15) | 4 (4.00) | 5 (21.74) | 0 (0.00) | 2 (11.11) | 0 (0.00) | |
| without accents | 168 (93.85) | 96 (96.00) | 18 (78.26) | 32 (100.00) | 16 (88.89) | 6 (100.00) | |
| 03_Articulation | N=178 | N=100 | N=23 | N=32 | N=17 | N=6 | 0.36 |
| legato | 167 (93.82) | 95 (95.00) | 20 (86.96) | 30 (93.75) | 17 (100.00) | 5 (83.33) | |
| staccato | 11 (6.18) | 5 (5.00) | 3 (13.04) | 2 (6.25) | 0 (0.00) | 1 (16.67) | |
| 04_Dynamic | N=177 | N=100 | N=23 | N=32 | N=16 | N=6 | 0.85 |
| f-ff | 19 (10.73) | 11 (11.00) | 3 (13.04) | 4 (12.50) | 1 (6.25) | 0 (0.00) | |
| mp-mf | 65 (36.72) | 37 (37.00) | 10 (43.48) | 12 (37.50) | 5 (31.25) | 1 (16.67) | |
| pp-mp | 93 (52.54) | 52 (52.00) | 10 (43.48) | 16 (50.00) | 10 (62.50) | 5 (83.33) | |
| 05_Interpretation | N=173 | N=97 | N=23 | N=32 | N=16 | N=5 | 0.39 |
| master performer | 130 (75.14) | 76 (78.35) | 15 (65.22) | 22 (68.75) | 12 (75.00) | 5 (100.00) | |
| mediocre performer | 43 (24.86) | 21 (21.65) | 8 (34.78) | 10 (31.25) | 4 (25.00) | 0 (0.00) | |
| 06_MelodicShape | N=170 | N=95 | N=23 | N=32 | N=15 | N=5 | 0.21 |
| jagged | 49 (28.82) | 25 (26.32) | 6 (26.09) | 14 (43.75) | 2 (13.33) | 2 (40.00) | |
| smooth | 121 (71.18) | 70 (73.68) | 17 (73.91) | 18 (56.25) | 13 (86.67) | 3 (60.00) | |
| 07_Meter | N=170 | N=95 | N=23 | N=32 | N=15 | N=5 | 0.70 |
| duple | 38 (22.35) | 23 (24.21) | 4 (17.39) | 9 (28.13) | 1 (6.67) | 1 (20.00) | |
| quadruple | 67 (39.41) | 33 (34.74) | 12 (52.17) | 13 (40.63) | 7 (46.67) | 2 (40.00) | |
| triple | 65 (38.24) | 39 (41.05) | 7 (30.43) | 10 (31.25) | 7 (46.67) | 2 (40.00) | |
| 08_RecordingQuality | N=166 | N=93 | N=21 | N=32 | N=15 | N=5 | 0.94 |
| high clarity | 143 (86.14) | 81 (87.10) | 17 (80.95) | 28 (87.50) | 13 (86.67) | 4 (80.00) | |
| low clarity | 23 (13.86) | 12 (12.90) | 4 (19.05) | 4 (12.50) | 2 (13.33) | 1 (20.00) | |
| 09_Repetitiveness | N=163 | N=93 | N=19 | N=32 | N=15 | N=4 | 0.77 |
| complete repetition | 87 (53.37) | 50 (53.76) | 8 (42.11) | 18 (56.25) | 8 (53.33) | 3 (75.00) | |
| no repetition | 76 (46.63) | 43 (46.24) | 11 (57.89) | 14 (43.75) | 7 (46.67) | 1 (25.00) | |
| 10_Register | N=161 | N=93 | N=17 | N=32 | N=15 | N=4 | 0.15 |
| high | 19 (11.80) | 8 (8.60) | 5 (29.41) | 6 (18.75) | 0 (0.00) | 0 (0.00) | |
| low | 41 (25.47) | 25 (26.88) | 4 (23.53) | 8 (25.00) | 4 (26.67) | 0 (0.00) | |
| middle | 101 (62.73) | 60 (64.52) | 8 (47.06) | 18 (56.25) | 11 (73.33) | 4 (100.00) | |
| 11_Rubato | N=161 | N=93 | N=17 | N=32 | N=15 | N=4 | 0.026 |
| with rubato | 129 (80.12) | 82 (88.17) | 12 (70.59) | 20 (62.50) | 12 (80.00) | 3 (75.00) | |
| without rubato | 32 (19.88) | 11 (11.83) | 5 (29.41) | 12 (37.50) | 3 (20.00) | 1 (25.00) | |
| 12_Tempo | N=159 | N=91 | N=17 | N=32 | N=15 | N=4 | 0.30 |
| fast | 56 (35.22) | 34 (37.36) | 8 (47.06) | 9 (28.13) | 3 (20.00) | 2 (50.00) | |
| medium | 69 (43.40) | 37 (40.66) | 9 (52.94) | 13 (40.63) | 9 (60.00) | 1 (25.00) | |
| slow | 34 (21.38) | 20 (21.98) | 0 (0.00) | 10 (31.25) | 3 (20.00) | 1 (25.00) | |

| | | | | | | | |
|---|---|---|---|---|---|---|---|
| 13_Texture | N=159 | N=91 | N=17 | N=32 | N=15 | N=4 | 0.56 |
| thick | 62 (38.99) | 32 (35.16) | 6 (35.29) | 16 (50.00) | 7 (46.67) | 1 (25.00) | |
| thin | 97 (61.01) | 59 (64.84) | 11 (64.71) | 16 (50.00) | 8 (53.33) | 3 (75.00) | |
| 14_Transition | N=158 | N=90 | N=17 | N=32 | N=15 | N=4 | 0.45 |
| continuous/smooth | 96 (60.76) | 54 (60.00) | 9 (52.94) | 22 (68.75) | 10 (66.67) | 1 (25.00) | |
| disjointed | 62 (39.24) | 36 (40.00) | 8 (47.06) | 10 (31.25) | 5 (33.33) | 3 (75.00) | |
| 15_Tonality | N=158 | N=90 | N=17 | N=32 | N=15 | N=4 | 0.31 |
| major | 76 (48.10) | 41 (45.56) | 8 (47.06) | 15 (46.88) | 8 (53.33) | 4 (100.00) | |
| minor | 82 (51.90) | 49 (54.44) | 9 (52.94) | 17 (53.13) | 7 (46.67) | 0 (0.00) | |
| 16_Familiarity | N=158 | N=90 | N=17 | N=32 | N=15 | N=4 | 0.83 |
| new excerpt | 37 (23.42) | 22 (24.44) | 2 (11.76) | 8 (25.00) | 4 (26.67) | 1 (25.00) | |
| previously heard excerpt | 121 (76.58) | 68 (75.56) | 15 (88.24) | 24 (75.00) | 11 (73.33) | 3 (75.00) | |

Data are presented as n (%). P-value was obtained from Chi-square test or Fisher's exact test.
